# Segmentation of vestibular schwannoma from MRI — An open annotated dataset and baseline algorithm

**DOI:** 10.1101/2021.08.04.21261588

**Authors:** Jonathan Shapey, Aaron Kujawa, Reuben Dorent, Guotai Wang, Alexis Dimitriadis, Diana Grishchuk, Ian Paddick, Neil Kitchen, Robert Bradford, Shakeel R Saeed, Sotirios Bisdas, Sébastien Ourselin, Tom Vercauteren

**Author notes:** corresponding author: Jonathan Shapey.

## Abstract

Automatic segmentation of vestibular schwannomas (VS) from magnetic resonance imaging (MRI) could significantly improve clinical workflow and assist patient management. We have previously developed a novel artificial intelligence framework based on a 2.5D convolutional neural network achieving excellent results equivalent to those achieved by an independent human annotator. Here, we provide the first publicly-available annotated imaging dataset of VS by releasing the data and annotations used in our prior work. This collection contains a labelled dataset of 484 MR images collected on 242 consecutive patients with a VS undergoing Gamma Knife Stereotactic Radiosurgery at a single institution. Data includes all segmentations and contours used in treatment planning and details of the administered dose. Implementation of our automated segmentation algorithm uses MONAI, a freely-available open-source framework for deep learning in healthcare imaging. These data will facilitate the development and validation of automated segmentation frameworks for VS and may also be used to develop other multi-modal algorithmic models.

## Background & Summary

Vestibular schwannoma (VS) is a benign tumour arising from the nerve sheath of one of the vestibular nerves. The incidence of VS was previously estimated to be between 14 and 20 cases per million per year^1^ but has risen significantly in recent years with approximately 1600 new patients now diagnosed every year in the UK.^2^. At current rates approximately 1 in 1000 people will be diagnosed with a VS in their lifetime^3,4^. As access to diagnostic Magnetic Resonance Imaging (MRI) has become more available, a greater number of asymptomatic patients with small tumours are being diagnosed with VS^1^. For smaller tumours, expectant management with lifelong imaging is often advised^5,6^. However, patients with larger tumours who undergo stereotactic radiosurgery (SRS) or conventional open surgery also require a prolonged period of surveillance after treatment.

In 2003, the international Consensus Meeting on Systems for Reporting Results in Vestibular Schwannoma agreed that the size of a VS should be defined by the tumour’s maximal linear dimension^6^. However, several studies have demonstrated that a volumetric measurement is a more accurate and sensitive method of calculating a vestibular schwannoma’s true size and is superior at detecting subtle growth^7,8^. Implementing routine volumetric measurements would therefore enable clinicians to offer earlier appropriate intervention. The principal reason such volumetric methods have not been widely implemented is because the currently available tools make calculating tumour volume assessment a labour-intensive process, prone to variability and subjectivity, with no dedicated software readily available within the clinical setting. An automated segmentation tool would also improve clinical workflow during the planning of SRS. It could for example be used as an initialisation step in the planning process, saving clinicians significant time, reducing cost and enabling more patients to be treated.

We have previously developed a novel artificial intelligence (AI) framework based on a 2.5D convolutional neural network (CNN) able to exploit the different in-plane and through-plane resolutions encountered in standard clinical imaging protocols^9,10^. In particular, we embedded a computational attention module to enable the CNN to focus on the small VS target and included a supervision on the attention map for more accurate segmentation. The method was trained and validated using contrast-enhanced T1-weighted and high-resolution T2-weighted images from a dataset of 242 consecutive patients treated with Gamma Knife SRS. Excellent accuracy scores were achieved (Mean Dice scores *>* 93%) comparable to repeated measurements performed by clinicians^10^.

In this work, we provide the first publicly-available imaging dataset of VS by releasing the data used in our prior work (Example dataset illustrated in figure 1). The release of this dataset and baseline segmentation code will facilitate the development and validation of automated segmentation frameworks for VS as well as support the development of novel applications. For example, more work is needed to enable our network to achieve a similar clinically-acceptable performance on general surveillance MRI scans rather than tightly controlled radiosurgery planning MRI protocols. The release of this dataset will support ongoing collaborative work in this area. This multi-modal imaging dataset will also facilitate algorithmic research relating to both multi-modal and modality-specific segmentation networks, to investigate methods of domain adaptation^11^ and to develop new methods of synthesising imaging data. Finally, by including each patient’s radiation treatment data, this dataset will also support work relating to the planning and delivery of radiation treatment including automated segmentation of organs at risk and automated dose planning.

**Figure 1.**
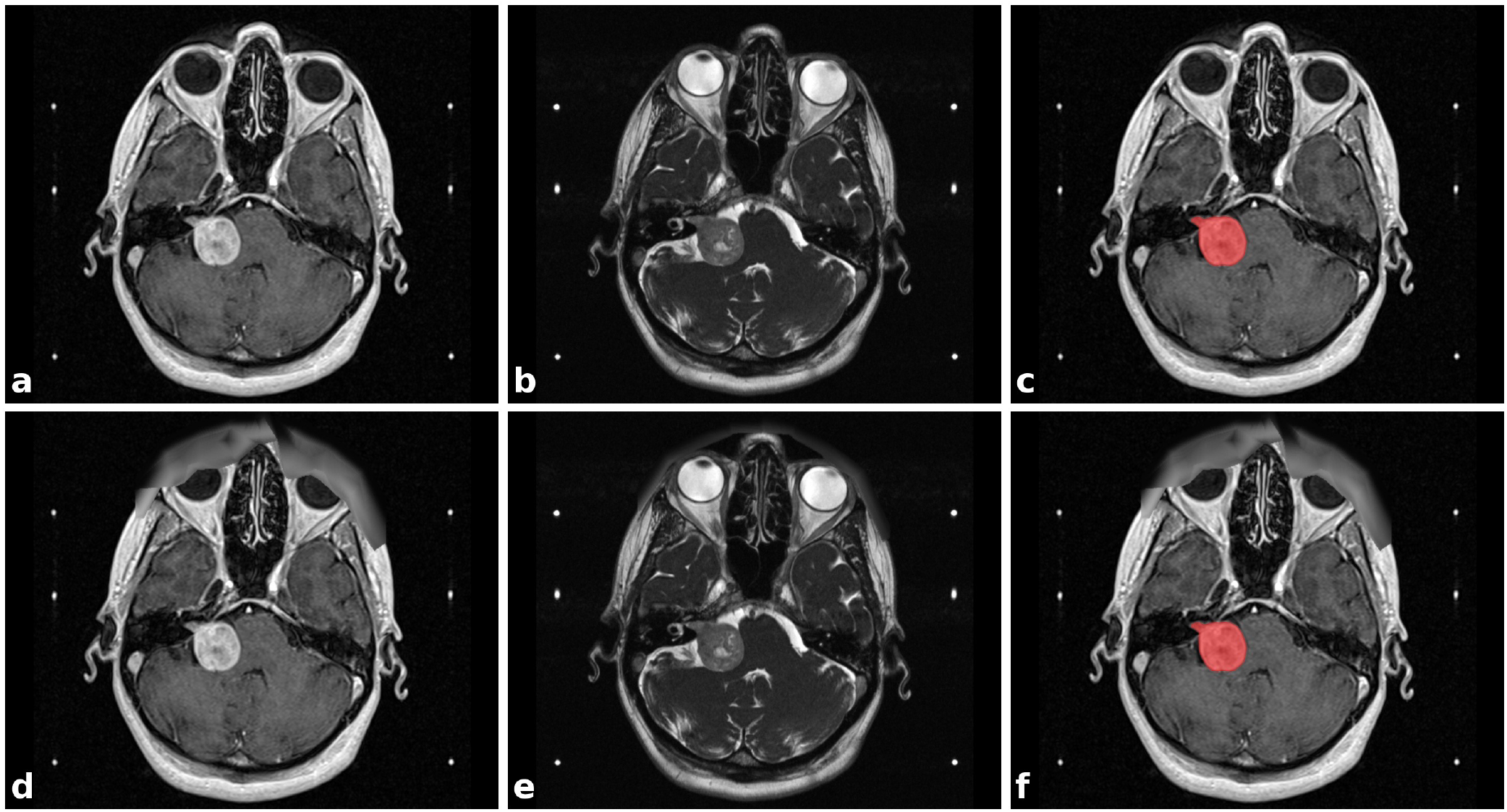
Illustrative example dataset of a patient with a right sided vestibular schwannoma (VS). a) Contrast-enhanced T1-weighted MRI (ceT1); b) High-resolution T2-weighted MRI (hrT2); c) ceT1 MRI with annotated segmentation of VS; d)-f) corresponding images after obscurification of facial features. The six white dots in each image are fiducials of the Leksell Stereotactic System MR Indicator box used for image co-registration.

## Methods

### Data overview

This collection contains a labelled dataset of 484 MR image sets collected on 242 consecutive patients (Male:Female 95:147; median age: 56 yr, range: 24 yr to 84 yr) with a VS undergoing Gamma Knife Stereotactic Radiosurgery (GK SRS) at a single institution. The treatment-planning MRI included in this dataset comprise contrast-enhanced T1-weighted (ceT1) images and high-resolution T2-weighted (hrT2) images acquired in a single MRI session prior to and typically on the day of the radiosurgery. Each imaging dataset is accompanied by the patient’s radiation therapy (RT) dataset including the RTDose, RTStructures and RTPlan DICOM files and the accompanying `contours.json` and `affines.tfm` files. All structures were manually segmented in consensus by the treating GK SRS neurosurgeon, neuroradiologist and physicist using both the ceT1 and hrT2 images.

Data were obtained from the Queen Square Radiosurgery Centre (Gamma Knife). Imaging data from consecutive patients with a single sporadic VS treated with GK SRS between October 2012 and January 2018 were screened for the study. All adult patients older than 18 years with a single unilateral VS treated with GK SRS were eligible for inclusion in the study, including patients who had previously undergone operative surgical treatment. In total, 248 patients met these initial inclusion criteria including 51 patients who had previously undergone surgery. None of the patients received prior radiation treatment. Patients were only included in the study if their pre-treatment image acquisition dataset was complete; 6 patients were thus excluded because of incomplete datasets. All details are included in the meta-data.

All contributions to this study were based on approval by the NHS Health Research Authority and Research Ethics Committee (18/LO/0532) and were conducted in accordance with the 1964 Declaration of Helsinki. Data were fully deidentified by removing all health information identifiers and by applying a de-facing algorithm as described hereafter. All data were visually inspected before release. Because patients were selected retrospectively and the MR images were completely de-identified before analysis, informed consent from individual participants was not required for the study. In addition, the terms of the data sharing agreements were approved by the Joint Research Office at University College London

### DICOM RT-objects

Three DICOM RT-objects were included for each MR image in this dataset : RT Structure Set, RT Plan and RT Dose. RT-objects were introduced in the DICOM standard to facilitate the exchange of specific information generated at different stages of the radiation therapy work flow. Specifically, these three RT-objects are created during the treatment planning phase based on the acquired images and using a treatment planning software. Each RT-object in this dataset refers to one MR image and its associated coordinate system.

The RT Structure Set defines areas of significance such as the VS on the day of the GammaKnife treatment. For some patients, organs at risk are also included, for example the cochlea. Each area is defined by a set of closed polygonal planar contour lines, which in turn are defined by the 3D-coordinates of their vertices. The RT Plan contains information about the treatment plan created for the patient, which includes the placement of the radiation beam, prescription and the patient setup. The RT Dose describes the predicted distribution of the radiation dose based on the treatment plan.

For a more detailed description of the RT-objects and their integration in the radiation work flow, the reader is referred to the introduction to the DICOM-RT standard provided by Law et al.^12^

### Image acquisition

All stereotactic images were obtained on a 32-channel Siemens Avanto 1.5T scanner using a Siemens single-channel head coil. Contrast-enhanced T1-weighted imaging was performed with an MPRAGE sequence with in-plane resolution of 0.4 *×* 0.4 mm, in-plane matrix of 512 *×* 512, and slice thickness of 1.0 mm to 1.5 mm (TR = 1900 ms, TE = 2.97 ms, TI = 1100 ms). High-resolution T2-weighted imaging was performed with a 3D CISS sequence in-plane resolution of 0.5 *×* 0.5 mm, in-plane matrix of 384 *×* 384 or 448 *×* 448, and slice thickness of 1.0 mm to 1.5 mm (TR = 9.4 ms, TE = 4.23 ms).

### Segmentation of tumours and other structures

Following acquisition of the stereotactic MRI scans, the image sequences were imported into the Gamma Knife planning software (Leksell GammaPlan software version 11.1, Elekta, Sweden). The stereotactic space was defined using the fiducial box coordinate system and once the image quality and stereotactic accuracy were verified the patient’s external contour (Skin) was created using the automatic image segmentation tool. The target tumour volume (TV) and organs at risk (OAR) such as the brainstem and cochlea were delineated by the treating GK SRS neurosurgeon, neuroradiologist and physicist. All segmentations (RTStructures) were performed using the GammaPlan software that employs an in-plane semi-automated segmentation method. Using this software, delineation was performed on sequential 2D axial slices to produce 3D models for each structure. Typically, the tumour was segmented on the ceT1 image and refined using the registered hrT2 image. In some cases where the tumour borders were better delineated on the hrT2, this image was used as the primary image for segmentation purposes. The cochlea was typically segmented on hrT2 image due to improved visualisation on this sequence.

The median target tumour volume (TV) within the dataset was 1.36 cm^3^ (range 0.04 *—* 10.78 cm^3^, IQR 0.63 *-*3.17 cm^3^). The dataset includes 49 post-surgical cases (20%).

### Details of radiation treatment

Once all segmentations were approved, a forward treatment planning approach was initiated by the medical physicist under the guidance of the treating clinician. Individual isocentres with varying positions, weightings and beam collimator configurations were placed into stereotactic space on different regions within the target. The median VS treatment dose was 13Gy (used in 211 87% of cases). 12Gy was used in 29 cases (12%) with 11Gy and 15Gy was used in one case each. The aim was to achieve adequate coverage of the TV by the prescription isodose (typically higher than 95%) whilst maintaining minimum spillage into surrounding tissue. Particular attention was placed to the anterior and medial surfaces of the target with the aim of achieving a high degree of conformity and steep dose fall-off. Attempts to reduce the dose to cochlea were also performed by blocking beam sectors that contributed dose to this OAR. The resulting treatment plan and isodose distributions were reviewed and manipulated until they were considered acceptable for delivery by the prescribing physician. Once the delivery platform passed the daily quality assurance checks, the treatment plan was exported to the treatment console for delivery.

### Data transmission and de-identification

De-identified data was submitted to The Cancer Imaging Archive (TCIA) through its established selection and governance processes^13^. To obscure facial features in the MR images, a face masking algorithm^14^ was applied to all images. Setting the T1 image as a reference for co-registration and a choosing grid step coefficient of 1.0 led to sufficient obscurification of facial features.

### Data Records

The images and associated files are publicly available on The Cancer Imaging Archive^15^ (TCIA: https://doi.org/10.7937/TCIA.9YTJ-5Q73)^16^. 4The archive data can be organized in the convenient folder structure shown in box 1 with a script from our code repository (https://github.com/KCL-BMEIS/VS_Seg). Alternatively, we provide a script to organize the archive data according to the Brain Imaging Data Structure (BIDS) specification.^17^

**Box 1.**
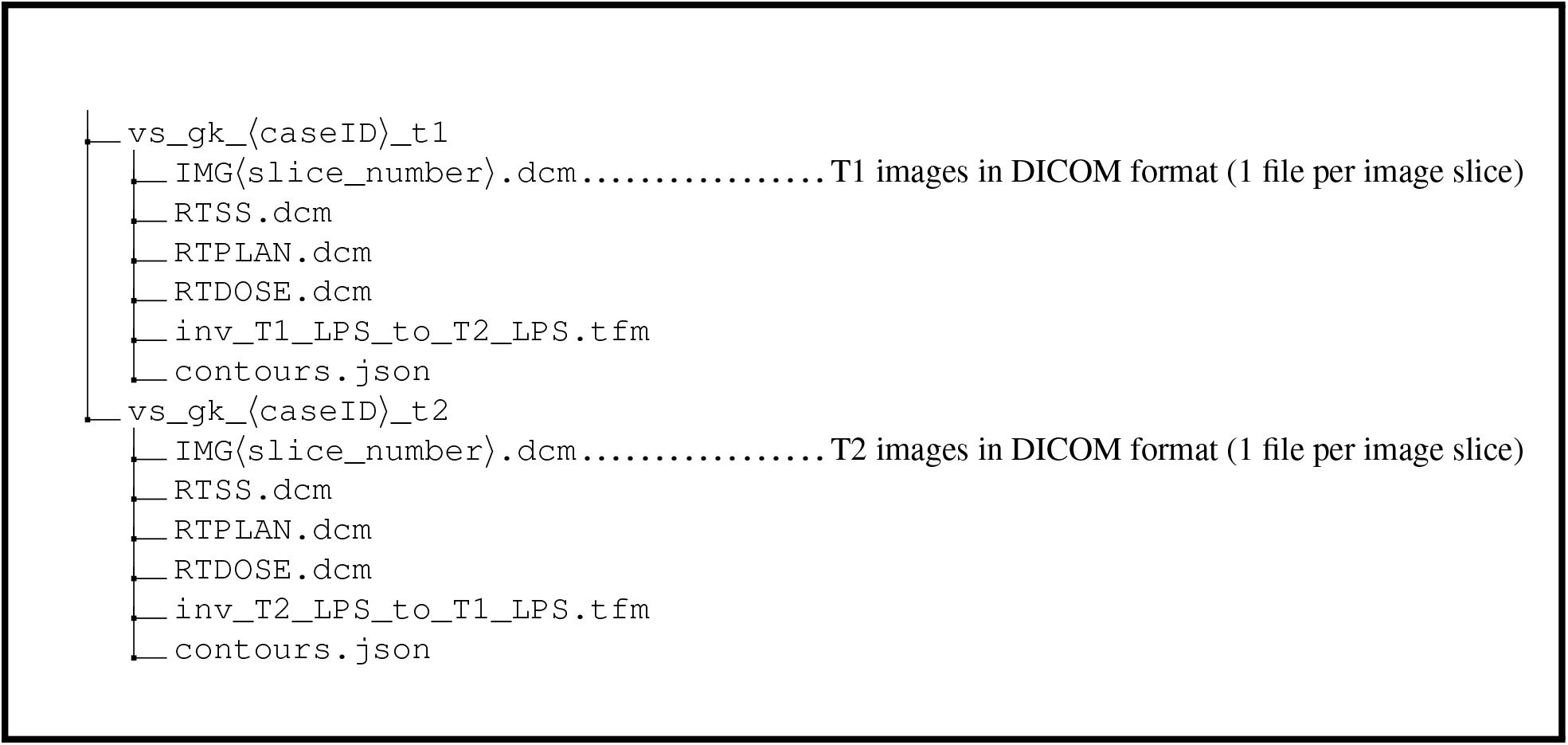
Folder structure of the data set

The VS image files are provided in DICOM format and the radiation therapy data in the DICOM-RT files `RTDOSE.dcm`, `RTPLAN.dcm`, and `RTSS.dcm`. The segmentation of the tumour is contained in the RTSS file.

The `.tfm` files are text files specifying affine transformation matrices that can be used to co-register the T1 image to the T2 image and vice versa. The file format is a standard format defined by the Insight Toolkit (ITK) library. The matrices are the result of the co-registration of fiducials of the Leksell Stereotactic System MR Indicator box into which the patient’s head is fixed during image acquisition. The localization of fiducials and co-registration is performed automatically by the Leksell GammaPlan software.

The two files named `contours.json` in the `T1` and `T2` folder contain the contour points of the segmented structures in JavaScript Object Notation (JSON) format, mapped in the coordinate frames of the T1 image and the T2 image, respectively.

In most cases, the tumour was segmented on the T1 image while the cochlea was typically segmented on the T2 image. This meant that some contour lines (typically for the tumour) were coplanar with the slices of the T1 image while others (typically for the cochlea) were coplanar with T2 slices. After co-registration, the (un-resampled) slices of the T1 and T2 image generally did not coincide, e.g. due to different image position and, occasionally, slice thickness.

Therefore, the combined co-registered contour lines were neither jointly coplanar with the T1 nor with the T2 image slices. This complicates the discretisation to a binary labelmap in either image space. Upon export of the segmentations in a given target space, the GammaPlan software interpolates between the original contour lines to create new slice-aligned contour lines in the target image space (T1 or T2). This results in the interpolated slice-aligned contour lines found in the `RTSS.dcm` files. In contrast, the contours in the JSON files were not interpolated after co-registration, and therefore describe the original (potentially off-target-space-slice) manual segmentation accurately.

Discretisation of the slice-aligned interpolated contour lines into a binary labelmap can be performed on a slice-by-slice basis with a simple 2D rasterisation algorithm. However, as shown in figure 2, when discretized to the target voxel grid space, starting from slice-aligned interpolated contours can result in unexpected behaviour such as the exclusion of entire slices of the segmentation at the top and bottom of a structure.

**Figure 2.**
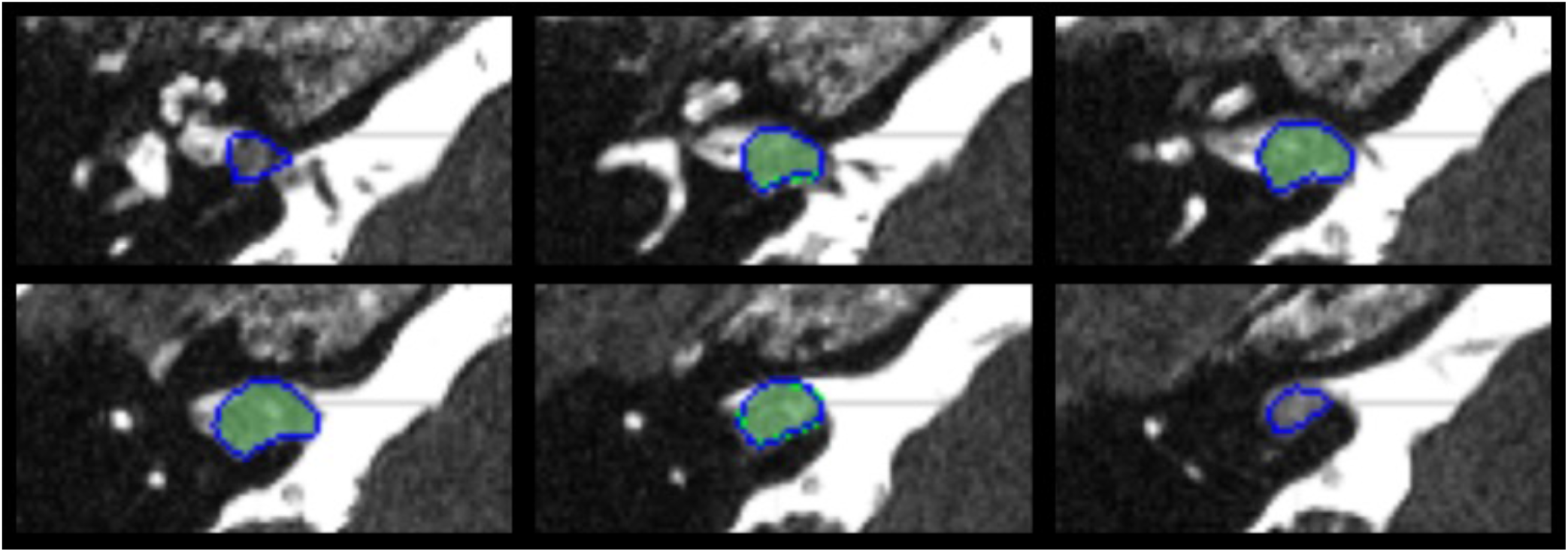
Comparison of the two contour discretization approaches. Both binary tumour labelmaps are obtained from original contours drawn on T1 slices overlaid on the (non-coinciding) T2 image slices of subject 182. Blue contours are the result of direct discretization in the T2 space from the original T1-aligned contours saved in the `JSON` files. Green labelmaps are the result of discretisation of the interpolated T2-slice-aligned contours saved in the `RTSS.dcm` files. In this worst-case example, the discretization of the interpolated contours results in a binary labelmap that misses the first and last slice compared to the labelmap obtained from the original contours. Intermediate slices are hardly affected by the interpolation and discretization strategy.

An alternative is to use an advanced discretization algorithm that can work directly from off-slices contours, such as that of Sunderland et al^18^. This algorithm is available in the open-source software 3D Slicer^19^ (www.slicer.org) with the extension SlicerRT^20^. It handles issues such as keyhole contours, rapid changes, and branching. We applied the same SlicerRT implementation of the algorithm of Sunderland et al^18^ to discretize both the slice-aligned interpolated contours and the original potentially off-slice contours. As it bypasses the proprietary interpolation performed by the closed-source GammaPlan software, starting from the original uninterpolated contour lines offers additional traceability of the discretisation process and offers a more transparent conversion. We observed that it results in a binary labelmap with a higher number of foreground pixels, as shown in figure 2. Due to the increased openness and consistency advantage, this approach was taken for the discretization of the manual segmentations into label maps used to train our automated segmentation algorithm.

### Technical Validation

The quality of the images in this dataset and their co-registration were assessed by radiologists during treatment planning. To verify the accuracy of the contours in the JSON file, they were compared to the contours stored in the RTSS files by converting both sets of contours into binary labelmaps using 3D Slicer, and calculating the Dice coefficient between them. The average Dice coefficient was 99.9*±* 0.2% for T1 images and 97.6*±* 2.2% for T2 images. All Dice coefficients were larger than 88%. The worst-case example is shown in figure 2. Any deviations from a perfect Dice coefficient of 100% are due to the interpolation of contours in the RTSS files.

Visual inspection was performed on all cases after applying the de-facing algorithm to ensure satisfactory obscurification of facial features.

For further validation, an automatic segmentation algorithm was trained and tested on the dataset. The comparison between predicted and manual segmentations showed high agreement with average Dice scores of 94.5 *±*2.2 % for T1 images and 90.7*±* 3.6 % for T2 images. This is comparable to the inter-observer variability between clinical annotators whose segmentations led to an average Dice score of 93.82 *±*3.08 %. Interobserver variability testing between clinical annotators recorded a Dice score of 93.82% (SD 3.08%), an ASSD score of 0.269 (SD 0.095) mm, and an RVE of 5.55% (SD 4.75%) between the 2 sets of manual annotations.

A comparison between the 2.5D UNet on which our implementation is based and a 3D UNet as well as other baseline neural networks without attention module and/or hardness-weighting was published in,^9^ showing improvements in Dice score of more than 3%.

### Usage Notes

To view the DICOM images we recommend 3D Slicer’s DICOM viewer (https://www.slicer.org), which is a free and open source platform for medical image informatics, image processing, and three-dimensional visualization. To load the RT Structure Set and overlay the image with the segmentation, the extension SlicerRT is required, which can be added through 3D Slicer’s extension manager. The `.tfm` files can also be opened with 3D Slicer or by directly using the ITK library. However, we also provide a script that performs the image registration and conversion from contours to binary labelmaps as described in the following section.

### Code availability

We have released a code repository for automated segmentation of VS with deep learning (https://github.com/KCL-BMEIS/VS_Seg). The applied neural network is based on the 2.5D UNet described in^9^ and^10^ and has been adapted to yield improved segmentation results. Our implementation uses MONAI, a freely available, PyTorch-based framework for deep learning in healthcare imaging (https://monai.io). This new implementation was devised to provide a starting point for researchers interested in automatic segmentation using state-of-the art deep learning frameworks for medical image processing.

To apply the algorithm to the VS dataset, we randomly split the final 242 patients into 3 non-overlapping groups: 176 for training, 20 for hyperparameter tuning, and 46 for testing, with median tumour volumes of 1.37 cm^3^ (range 0.04 − 9.59 cm^3^, IQR 0.64 − 3.14 cm^3^), 0.92 cm^3^ (range 0.12 − 5.50, IQR 0.51 − 2.40 cm^3^), and 1.89 cm^3^ (range 0.22 − 10.78, IQR 0.74 − 4.05 cm^3^), respectively. Thirty-four patients (19%) in the training dataset had undergone previous surgery compared with 2 patients (2%) in the hyperparameter tuning set and 14 patients (30%) in the testing dataset. The algorithm leads to excellent average Dice scores between predicted segmentations and ground truth (see Technical Validation). To apply the released code to the VS data set, the DICOM images and RT Structures have to be converted into NIfTI files. The repository includes a script and instructions for this purpose.

## Data Availability

The images and associated files are publicly available on The Cancer Imaging Archive (TCIA).

https://doi.org/10.7937/TCIA.9YTJ-5Q73

## Acknowledgements

This work was supported by Wellcome Trust (203145Z/16/Z, 203148/Z/16/Z, WT106882), EPSRC (NS/A000050/1, NS/A000049/1) and MRC (MC/PC/180520) funding. Tom Vercauteren is also supported by a Medtronic/Royal Academy of Engineering Research Chair (RCSRF1819/7/34)..

## Author contributions statement

J.S. conceived the research. N.K., R.B., I.P. and A.D. were the treating clinicians/physicists. J.S., A.D. and D.G. completed the data collection. A.K., R.D. and G.W. devised and implemented the code. J.S., A.K, R.D., G.W. and T.V. analysed the data. All authors reviewed the manuscript.

